# An exact method for quantifying the reliability of end-of-epidemic declarations in real time

**DOI:** 10.1101/2020.07.13.20152082

**Authors:** Kris V Parag, Christl A Donnelly, Rahul Jha, Robin N Thompson

**Affiliations:** MRC Centre for Global Infectious Disease Analysis, Imperial College London, London, W2 1PG, UK; Department of Statistics, University of Oxford, Oxford, OX1 3LB, UK; Department of Applied Math and Theoretical Physics, University of Cambridge, Cambridge CB3 0WA, UK; Mathematical Institute, University of Oxford, Oxford, OX2 6GG, UK

**Keywords:** epidemic elimination, renewal processes, reproduction numbers, epidemic curves, Bayesian statistics, infectious disease, second waves, epidemic extinction

## Abstract

We derive and validate a novel and analytic method for estimating the probability that an epidemic has been eliminated (i.e. that no future local cases will emerge) in real time. When this probability crosses 0.95 an outbreak can be declared over with 95% confidence. Our method is easy to compute, only requires knowledge of the incidence curve and the serial interval distribution, and evaluates the statistical lifetime of the outbreak of interest. Using this approach, we rigorously show how the time-varying under-reporting of infected cases will artificially inflate the inferred probability of elimination, leading to premature (false-positive) end-of-epidemic declarations. Contrastingly, we prove that incorrectly identifying imported cases as local will deceptively decrease this probability, resulting in delayed (false-negative) declarations. Failing to sustain intensive surveillance during the later phases of an epidemic can therefore substantially mislead policymakers on when it is safe to remove travel bans or relax quarantine and social distancing advisories. World Health Organisation guidelines recommend fixed (though disease-specific) waiting times for end-of-epidemic declarations that cannot accommodate these variations. Consequently, there is an unequivocal need for more active and specialised metrics for reliably identifying the conclusion of an epidemic.

**Author Summary:** Deciding on when to declare an infectious disease epidemic over is an important and non-trivial problem. Early declarations can mean that interventions such as lockdowns, social distancing advisories and travel bans are relaxed prematurely, elevating the risk of additional waves of the disease. Late declarations can unnecessarily delay the re-opening of key economic sectors, for example trade, tourism and agriculture, potentially resulting in significant financial and livelihood losses. Here we develop and test a novel and exact data-driven method for optimising the timing of end-of-epidemic declarations. Our approach converts observations of infected cases up to any given time into a prediction of the likelihood that the epidemic is over at that time. Using this method, we quantify the reliability of end-of-epidemic declarations in real time, under ideal case surveillance, showing that it can depend strongly on past infection numbers. We then prove that failing to compensate for practical issues such as the time-varying under-reporting and importing of cases necessarily results in premature and delayed declarations, respectively. These variations and biases cannot be accommodated by current worldwide declaration guidelines. Sustained and intensive surveillance coupled with more adaptive declaration metrics are vital if informed end-of-epidemic declarations are to be made.

## Introduction

The timing of an end-of-epidemic declaration can have significant economic and public health consequences. Early or premature declarations can negate the benefits of prior control measures (e.g. quarantines or lockdown), leaving a population at an elevated risk to the resurgence of the infectious disease. The Ebola virus epidemic in Liberia (2014-2016), for example, featured several declarations that were followed by additional waves of infections [1]. Late or delayed declarations, however, can unnecessarily stifle commercial sectors such as agriculture, trade and tourism, leading to notable financial and livelihood losses. One of the first studies advocating the need for improved end-of-epidemic metrics suggested that the MERS-CoV epidemic in South Korea was declared over at least one week later than was necessary [2]. Balancing the health risk of a second wave of infections against the benefits of reopening the economy earlier is a non-trivial problem and is currently of major global concern as many countries prepare to meet the challenge of resurging COVID-19 caseloads.

World Health Organisation (WHO) guidelines adopt a time-triggered (i.e. decisions are enacted after some fixed, deterministic time) approach to end-of-epidemic declarations, recommending that officials wait for some prescribed period after the last observed infected case has recovered, before adjudging the outbreak over. The most common waiting time, which applies to Ebola virus and MERS-CoV among others, involves twice the maximum incubation period of the disease [3]. While having a fixed decision time is simple and actionable, it neglects the stochastic variation that is inherently possible at the tail of an outbreak. Recent studies have started to question this time-triggered heuristic and to investigate the factors that could limit its practical reliability.

Initial advances in this direction were made in [2], where mathematical formulae for assessing the end of an epidemic, in a data-driven manner, were derived. These formulae use the time-series of new cases (incidence) across an epidemic together with estimates of its serial interval distribution and basic reproduction number to compute the probability that the outbreak is over at any moment. The serial interval distribution describes the random inter-event times between the onset of symptoms of an infector and infectee, while the basic reproduction number is the average number of secondary infections per primary infection at the start of an epidemic [4, 5]. The output of this method is an epidemiologically informed statistical measure of confidence in an end-of-epidemic declaration.

This approach is important, but not perfect. It assumes that infected cases are reported without any error and it depends on parameters that relate to the initial growth phase of the epidemic. Moreover, to maintain simplicity, it adopts a mathematically conservative description of transmission, making its end-of-epidemic declaration time estimates likely to be late or delayed [2]. More recent studies [6, 7] have applied forward simulation to investigate the tail dynamics of an outbreak. These have revealed the impact of the constant under-reporting of cases [6] and demonstrated the sensitivity of declarations to the effective reproduction number [7], a parameter that generalises the basic reproduction number and that remains relevant across all phases of the epidemic. The influence of different routes of transmission on declarations has also been examined in [1] using the framework of [2].

However, there is still much we do not know about the dynamics of an outbreak as it approaches its end. Specifically, analytic and general insight into the sensitivity of end-of-epidemic declarations to practical surveillance imperfections is needed. Real incidence data is corrupted by time-varying trends in under-reporting, delays in case notification and influenced by the interaction of imported and local cases [8, 9, 10]. Previous works have either assumed perfect reporting [2] or treated constant under-reporting within some simulated scenarios [6, 7]. Here we attempt to expose the implications of more realistic types of data corruption, particularly time-varying case under-reporting and importation, by developing an exact framework that provides broad and provable insights. Understanding how realistic surveillance patterns can bias our perception of the epidemic end is the first step to engineering sensible and effective countermeasures against these biases.

We build on the renewal process transmission model from [11, 12], to derive and test a novel and exact real-time method for estimating the probability of elimination; defined as the probability that no future local cases will emerge conditioned on the past epidemic incidence. We explain this model in Fig. 1. Using this probability, we define an event-triggered [13, 14] declaration metric that guarantees confidence in that declaration provided the assumptions of the model hold. The trigger is the first time that this probability crosses a threshold e.g. we are 95% confident in our declaration if the threshold is 0.95. Event-triggered decision-making was essentially proposed by [2], has proven effective in other fields [15, 16, 17] and belies the time-triggered WHO approach, which fixes the time (elapsed since the last case^1^) but not the confidence in declaration.

**Fig. 1:**
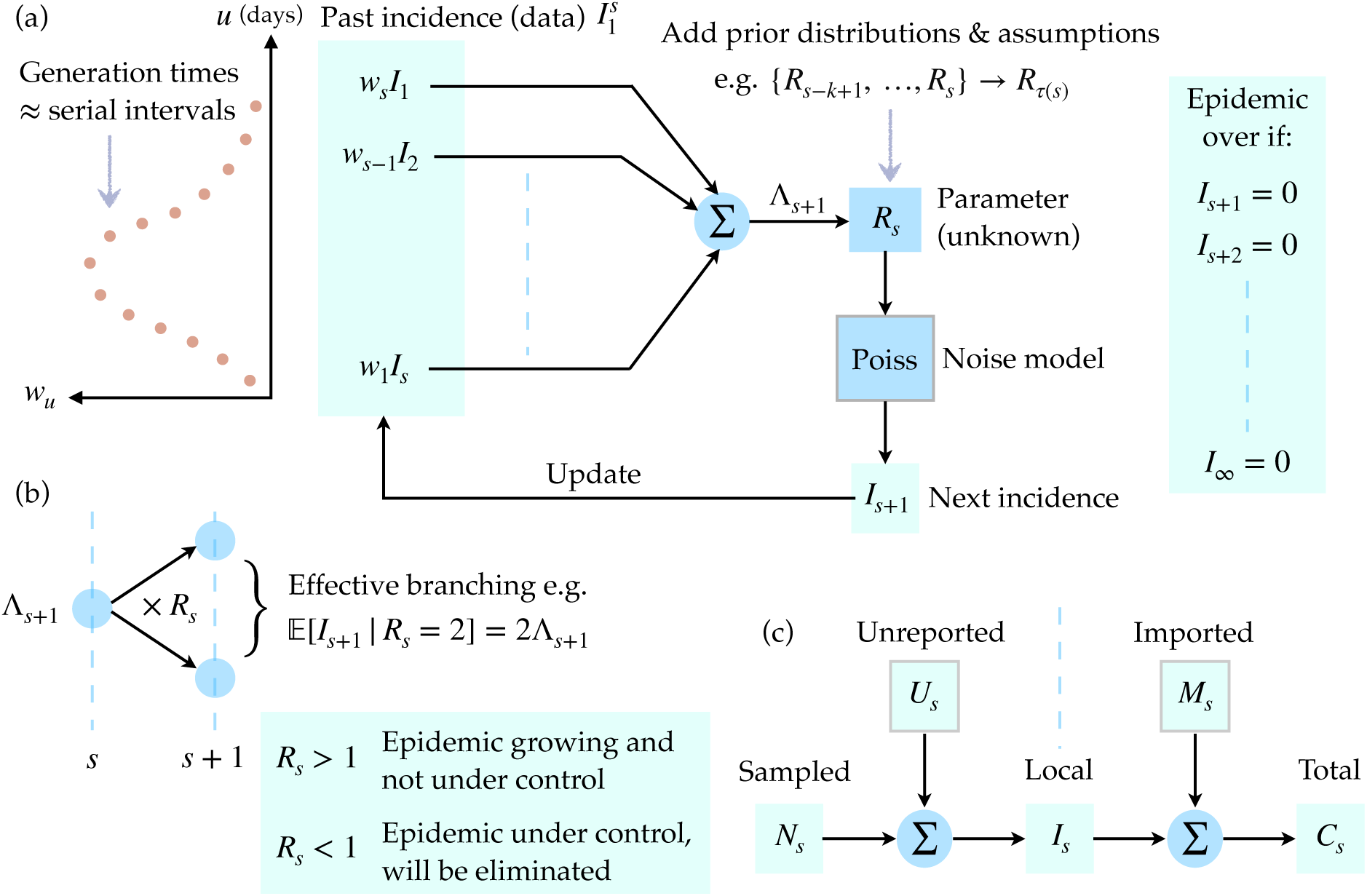
Transmission dynamics of an infectious disease. The renewal approach to infection propagation is outlined under a Poisson noise model in panel (a). Past, observed infected cases 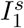, which form an incidence curve, seed new infections with probabilities proportional to *w*_*u*_ defined by the generation time distribution of the disease, which is approximated by the serial interval distribution. The total infectiousness Λ_*s*+1_ sums the contributions of past cases according to the set of {*w*_*u*_}. The effective reproduction number *R*_*s*_ determines how many effective infections are passed on to the next time unit *s* + 1. It is common to group *R*_*s*_ values over a window *τ* (*s*) to improve estimation reliability. When all future incidence values are zero we conclude that the epidemic is over or eliminated. Panel (b) shows how *R*_*s*_ acts as a reproductive parameter, controlling whether the epidemic grows or dies out. This parameter is therefore essential to predicting the dynamics of an epidemic. Panel (c) provides a breakdown of more realistic observation assumptions, where we might not be able to directly measure the local and complete incidence *I*_*s*_ due to unreported *U*_*s*_ or imported (migrating) *M*_*s*_ cases. If we can only observe sampled cases, *N*_*s*_, or the total number of cases, *C*_*s*_, then our epidemic predictions will be biased.

We benchmark our estimate against the true probability of elimination, i.e. the probability if the statistics and effective reproduction number of the epidemic were known precisely, and show consistency under the perfect conditions in [2] but with the caveat that we estimate effective reproduction numbers from the incidence curve in real time. We find that even the true elimination probabilities strongly depend on the specific stochastic incidence curve observed, confirming that time-triggered decision heuristics are unwarranted. Using our exact framework we prove two key results about imperfect surveillance. First, any type of time-varying under-reporting will lead to premature or false-positive event-triggers and hence declarations, unless explicit knowledge of the under-reporting scheme is available. Second, a failure to identify and account for the differences between local and imported cases will result in delayed or false-negative event-triggers, regardless of the dynamics of case importation.

Many infectious disease epidemics, including the ongoing COVID-19 pandemic, are known to feature extensive time-varying under-reporting and repeated importations from different regions [18, 19]. As this pandemic progresses into a potential second wave in several countries, public health authorities will need to decide when to relax and reapply intervention measures such as lockdowns, social distancing policies or travel bans [20]. Our work suggests that intensive surveillance, both of cases and their origin, must be sustained to make informed, reliable and adaptive decisions about the threat posed by the virus in the waning stages of the outbreak, even if reported case numbers remain at zero for consecutive days. We hope that our method, which is available at https://github.com/kpzoo/End-of-epidemic-declarations, will aid understanding and assessment of the tail kinetics of infectious epidemics.

## I. Methods

### Infectious disease transmission models

We can mathematically describe the transmission of an infection within a population over time with a renewal process based on the Euler-Lotka equation from ecology and demography [4]. This process models communicable pathogen spread from a primary (infected) case to secondary ones at some time *s* using two key variables: the effective reproduction number, *R*_*s*_, and the generation time distribution with probabilities {*w*_*u*_} for all times *u*. Here *R*_*s*_ defines the number of secondary cases at time *s* + 1 one primary case at *s* infects on average, while *w*_*u*_ is the probability that it takes *u* time units for a primary case to infect a secondary one [4]. As infection events are generally unobserved, we approximate the time of primary and secondary infection with the corresponding times of symptom onset i.e. the serial interval. This amounts to making the common assumption that the serial interval distribution, which can be observed, is a good approximation to the generation time distribution [2, 12].

If *I*_*s*_ counts the newly observed infected cases at *s* and a Poisson (Poiss) model is used to represent the noise in these observations then the renewal model captures the reproductive dynamics of infectious disease transmission with *I*_*s*_ ∼ Poiss(*R*_*s*−1_Λ_*s*_) [5]. Here 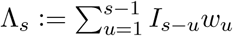 is the total infectiousness of the disease up to time *s* − 1 and summarises how previous cases contribute to upcoming cases on day *s*. We use 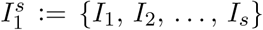 to represent the incidence curve from time 1 to *s*. A schematic of this approach to epidemic transmission is given in Fig. 1. Usually we are interested in estimating the *R*_*s*_ numbers in real time [21, 22] from the progressing 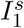, assuming that the serial interval distribution is known (i.e. derived from some other linelist data) [12].

This effective reproduction number is important for forecasting the kinetics of the epidemic. If *R*_*s*_ *>* 1 then we can expect the number of infections to increase monotonically with time. However, if *R*_*s*_ *<* 1 is sustained then we can be confident that the epidemic is being controlled and will, eventually, be eliminated [23]. In order to enhance the reliability of these estimates we usually assume that the epidemic transmission properties are stable over a look-back window of size *k* defined at time *s* as *τ* (*s*) := {*s, s*−1, …, *s*−*k*+1} [12, 24]. *We let the reproduction number over this window be R*_*τ* (*s*)_ and apply a conjugate gamma (Gam) prior distribution assumption: *R*_*τ* (*s*)_ ∼ Gam (*a*, 1*/c*) with *a* and *c* as shape-scale hyperparameters. This formulation, together with the use of gamma prior distributions, is standard in current renewal model frameworks [12, 21, 25].

The posterior distribution of *R*_*τ* (*s*)_ given the relevant window of the past incidence curve of data i.e. 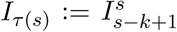 is also gamma distributed as follows [22]

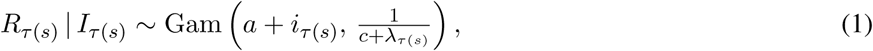

with grouped sums *i*_*τ* (*s*)_ := Σ_*u*∈*τ* (*s*)_ *I*_*u*_ and *λ*_*τ* (*s*)_ := Σ_*u*∈*τ* (*s*)_ Λ_*u*_. If some variable *y* ∼ Gam(*α, β*) then ℙ(*y*) = *y*^*α*−1^*e*^−*y/β*^*/β*^*α*^Γ(*α*) and 𝔼[*y*] = *αβ*. As a result, Eq. (1) yields the posterior mean estimate of 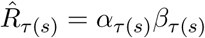 with *α*_*τ*(*s*)_ := *a* + *i*_*τ*(*s*)_, *β*_*τ*(*s*)_ := 1*/c*+*λ*_*τ* (*s*)_. Eq. (1) allows us to infer the grouped or averaged effective reproduction number over the window *τ* (*s*), which is considered an approximation of the unknown *R*_*s*_.

We can derive the posterior predictive distribution of the next incidence value (at time *s* + 1) by marginalizing over the domain of *R*_*τ* (*s*)_ as in [22]. If the space of possible predictions at *s* + 1 is *x* | *I*_*τ* (*s*)_ and NB indicates a negative binomial distribution then we obtain

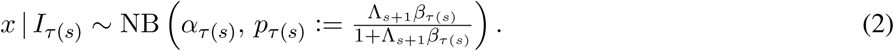

Eq. (2) completely describes the uncertainty surrounding one-step-ahead incidence predictions and is causal because all of its terms (including Λ_*s*+1_) only depend on the past observed incidence curve 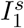 [22].

If a random variable *y* ∼ NB(*α, p*) then 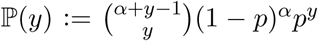 and 𝔼[*y*] = *pα/*1−*p*. Hence our posterior mean prediction is 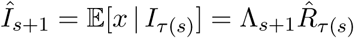. The current estimate of *R*_*τ* (*s*)_ influences our ability to predict upcoming incidence points. Thus, we expect that good estimation of the effective reproduction number is necessary for projecting the future behaviour of an infectious disease epidemic. In Results we rigorously extend and apply this insight to derive an exact method for computing the probability that an epidemic is reliably over at some time *s* i.e. that no future infections will occur from *s* + 1 onwards.

### Under-reported and imported cases

The above formulation assumes perfect case reporting and that all cases, 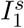, are local to the region being monitored. We now relax these assumptions. First, we consider more realistic scenarios where only some fraction of the local cases are reported or observed at any time. We use *N*_*s*_ and *U*_*s*_ for the number of sampled and unreported cases at time *s*. We consider a general time-varying binomial (Bin) sampling model with 0 ≤ *ρ*_*s*_ ≤ 1 as the probability that a true case is sampled at time *s* (hence 1 − *ρ*_*s*_ is the under-reporting probability). Then *N*_*s*_ ∼ Bin(*I*_*s*_, *ρ*_*s*_). The smaller *ρ*_*s*_ is, the less representative the sampled curve 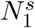 is of the true 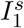.

This is a standard model for under-reporting [8, 26] and implies the following statistical relationship

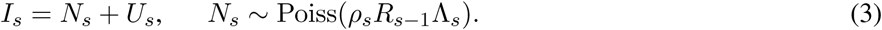

Raikov’s theorem [27] states that if the sum of two independent variables is Poisson then each variable is also Poisson. Consequently, *U*_*s*_ is Poisson with mean (1 − *ρ*_*s*_)*R*_*s*−1_Λ_*s*_. Most studies assume that *ρ*_*s*_ = *ρ* for all *s* i.e. that constant under-reporting occurs. The persistence of the Poisson relationship in Eq. (3) means that we can directly apply the forecasting and estimation results of the previous section to *N*_*s*_. Practically, if we observe only 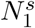 then unless we have independent knowledge of *ρ*_*s*_ (which can often be difficult to ascertain reliably [18, 26]) we can only construct an approximation to *ρ*_*s*_Λ_*s*_ as 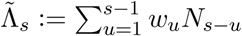 with 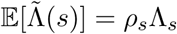.

Second, we investigate when imported or migrating cases from other regions, denoted by count *M*_*s*_ at time *s*, are introduced, resulting in the total number of observed cases being *C*_*s*_. Within this framework we ignore the under-reporting of cases and assume that *I*_*s*_ is observed to avoid confounding factors. We follow the approach of [9] and describe *M*_*s*_ as a Poisson number with some mean at time *s* of *ϵ*_*s*_. Using Raikov’s theorem we obtain

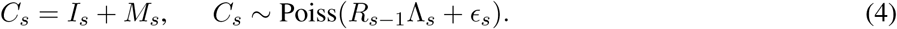

Eq. (4) models how imported cases combine with existing local ones to propagate future local infections.

While our work does not require assumptions on *ϵ*_*s*_, for ease of comparison later on we adopt the convention that the sum of imports and local cases drive the epidemic forward with the same reproduction number and serial interval [28]. Consequently, 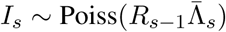 with 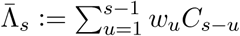. Practically, when surveillance is poor (i.e. local and imported cases cannot be distinguished), it is common to assume that all observed cases are local and conform to the approximate model 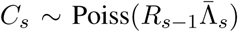 [25]. The forecasting and estimation results of the previous section therefore also apply under these conditions.

In Results we examine the impact of imperfect (our null hypothesis ℋ_0_) and ideal (the alternative ℋ_1_) surveillance within the context of under-reporting and importation in turn. We treat each problem individually to isolate the impact of each bias. Ideal surveillance then represents the ability to know either *U*_*s*_ or *M*_*s*_ (depending on the problem of interest) and hence account for their contributions. Imperfect surveillance refers to only having knowledge of *N*_*s*_ or *C*_*s*_ and basing inferences on these curves under the strong assumption that they approximate the true incidence. This assumption is often made in the literature [2, 12, 21] for the purposes of tractability and means Eq. (1) and Eq. (2) are valid. Fig. 1 summarises the relationships from Eq. (3) and Eq. (4).

## Results

### An exact method for declaring an outbreak over

We define an epidemic to be eliminated or over [23] at time *s* if no future, local or indigenous infected cases are observed i.e. *I*_*s*+1_ = *I*_*s*+2_ = *…* = *I*_∞_ = 0. We can define the estimated probability of elimination, *z*_*s*_, as

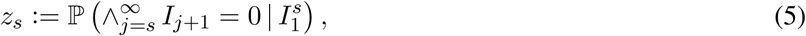

with 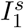 as the incidence curve (data), observed until time *s*. We refer to *z*_*s*_ as an estimated probability because we do not have perfect knowledge of the epidemic statistics e.g. we cannot know *R*_*s*_ precisely. The importance of this distinction will become clear in the subsequent section (see Eq. (10)). However, we observe that if we could have this idealised knowledge then Eq. (5) would exactly define the probability of no future cases given 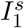.

Declaring the end of an epidemic with confidence *µ*% translates into solving the optimal stopping time problem

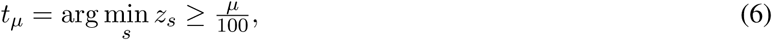

with *t*_95_, for example, signifying the first time that we are at least 95% sure that the epidemic has ended. Note that *z*_*s*_ is a function of 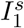 and practically characterises our uncertainty in the outcome of the epidemic (i.e. if it is over or not). This uncertainty derives from the fact that a range of possible epidemics with distinct future incidences 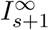 can possess the same 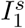 and 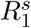 values. Some uncertainty exists even if 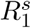 is known perfectly.

Eq. (6) presents an event-triggered approach to declaring the end of an epidemic with the *µ* threshold serving as an informative trigger. Event-triggered formulations have the advantage of being robust to changes in the observed data [13, 14], a point visible from the dependence of *z*_*s*_ and hence *t*_*µ*_ on 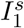. While Eq. (6) is written in absolute time, we may also clock time relative to the last observed case, *t*_0_. Our waiting time until declaration is then Δ*t*_*µ*_ = *t*_*µ*_ − *t*_0_, which is more useful for comparing *z*_*s*_ values from various realisations of 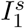 and for deriving confidence intervals. Later, we consider differences in the Δ*t*_*µ*_, denoted *δt*_*µ*_, proposed by comparable methods.

Previous works on end-of-epidemic declarations have either approximated *z*_*s*_ with a simpler, more conservative probability [2] or used simulations to estimate a quantity similar to *z*_*s*_ that is averaged over those simulations [6] [7]. No study has yet (to our knowledge) included real-time estimates of *R*_*s*_, within its assessment of epidemic elimination, despite the importance of this parameter in foretelling transmission [23]. By taking the renewal process approach to epidemic propagation (see Fig. 1), we explicitly embed uncertainty about *R*_*s*_ estimates to obtain an analytic and insightful expression for the probability that the outbreak is over given past observed cases (Eq. (5)). We derive this by inferring *R*_*s*_ within a sequential Bayesian framework from 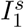, using a moving window of length *k* time units. We denote this estimate *R*_*τ* (*s*)_ with window *τ* (*s*) spanning 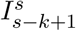 [12, 22]. Our main result is summarised as a theorem below (see Methods for further details and notation). Fig. 2 illustrates how our computed *z*_*s*_ probability varies across the lifetime of an example incidence curve, thus providing a real-time, causal and dynamically updating view of our confidence in its end.

**Fig. 2:**
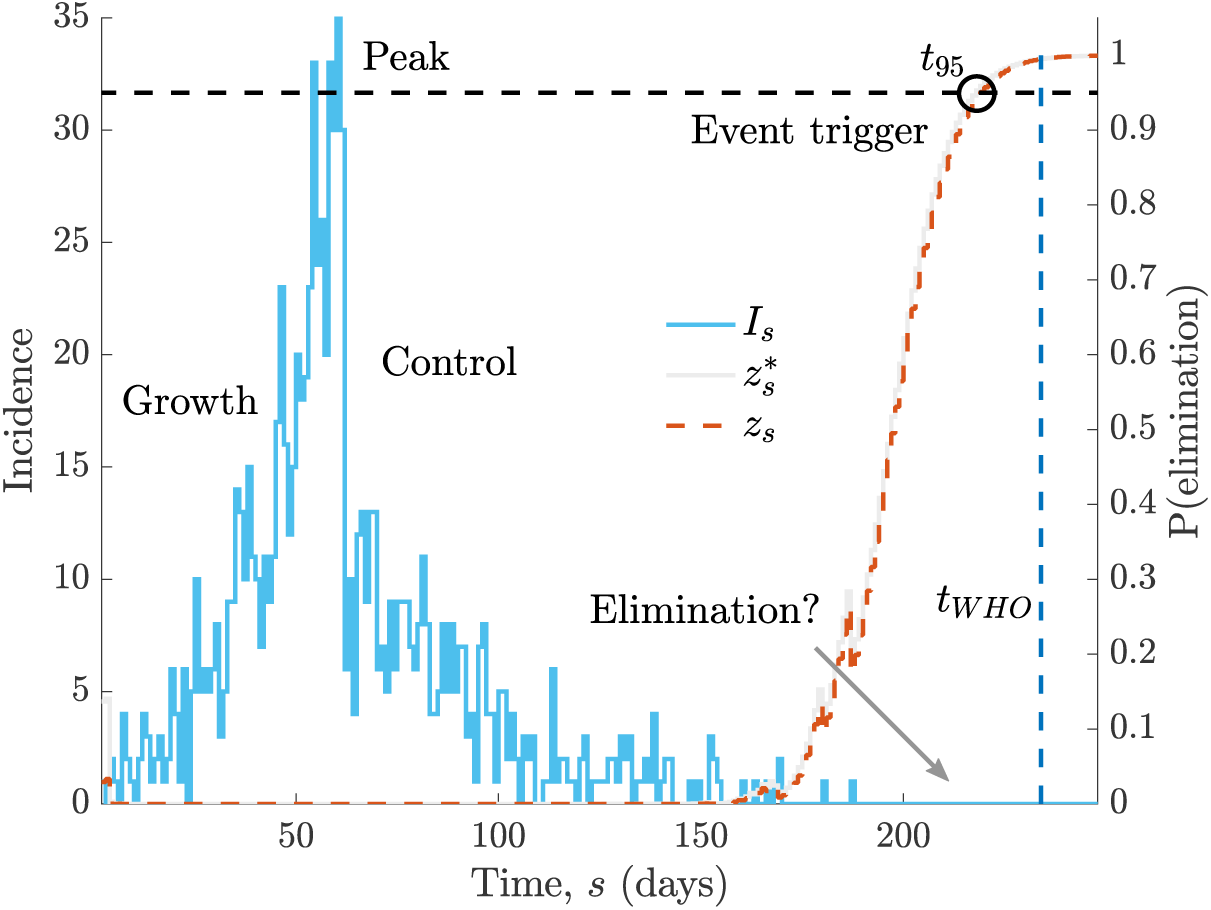
Elimination probabilities across the lifetime of an epidemic. We simulate a single incidence curve, *I*_*s*_ (blue, case counts on left y-axis), under the serial interval distribution for Ebola virus [29] and a true *R*_*s*_ profile that step changes from 2 to 0.5 at *s* = 100 days. We compute the true and estimated elimination probabilities, 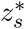 and *z*_*s*_, conditional on all cases observed up to time *s* in grey and red respectively (right y-axis). The circle (black) indicates when the outbreak can be declared over with 95% confidence. Observe how *z*_*s*_ and 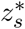 respond to the low *I*_*s*_ at the beginning of the epidemic before remaining 0 until we get to the tail of the outbreak, where a couple fluctuations occur due to some final cases. An estimate of the WHO declaration time, *t*_*WHO*_ [3], which is mostly insensitive to past case profiles is in dark blue. The central question in this study is how few cases need to be observed in the recent past before we can be confident that the epidemic has been eliminated.

#### Theorem 1.

If the posterior distribution of the grouped effective reproduction number, *R*_*τ* (*s*)_, given the incidence curve 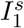 has form Gam(*α*_*τ* (*s*)_, *β*_*τ* (*s*)_) then the estimated probability that this epidemic has been eliminated at time *s* is 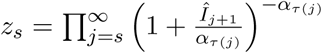 with 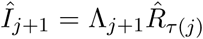 and 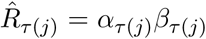 as the mean posterior incidence prediction and effective reproduction number estimate at time *j*, respectively.

We outline the development of this theorem. First, we decompose Eq. (5) into sequentially predictive terms as:

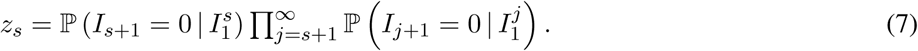

For simplicity, we rewrite Eq. (7) as 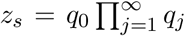. The factor *q*_*j*_ conditions on 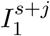, which includes all the epidemic data, 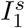 and the sequence of assumed zeros beyond that i.e. 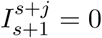 for *j* ≥ 1. This sequence is treated as pseudo-data. Observe that *q*_0_ is simply a one-step-ahead prediction of 0 from the available incidence curve.

We solve Eq. (7) by making use of known renewal model results derived in [12, 22, 24] and outlined in Methods. The renewal transmission model allows us to estimate the effective reproduction number *R*_*s*_ and hence compute *z*_*s*_ in real time (see Fig. 1). This estimate at time *s, R*_*τ* (*s*)_, uses the look-back window *τ* (*s*) of *k* time units (e.g. days). The posterior over *R*_*τ* (*s*)_ is shape-scale gamma distributed as Gam(*α*_*τ* (*s*)_, *β*_*τ* (*s*)_) with *α*_*τ* (*s*)_ := *a* + *i*_*τ* (*s*)_ and 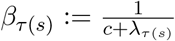 (see Eq. (1)). Here (*a, c*) are hyperparameters of a gamma prior distribution placed on *R*_*τ* (*s*)_ and *i*_*τ* (*s*)_ and *λ*_*τ* (*s*)_ are grouped sums of the incidence *I*_*u*_ and total infectiousness Λ_*u*_ for *u* ∈ *τ* (*s*). The total infectiousness describes the cumulative impact of past cases and is defined in Methods.

Under this formulation, the posterior predictive distribution of the incidence at *s* + 1 is negative binomially distributed (NB) (see Eq. (2)). The probability of *I*_*s*+1_ being zero from this distribution gives 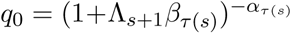 by substitution. The next term, *q*_1_, is computed similarly because we condition on *I*_*s*+1_ = 0 as pseudo-data (i.e. the sequential terms in Eq. (7)) and update Λ_*s*+2_, *β*_*τ* (*s*+1)_ and *α*_*τ* (*s*+1)_ with this zero. Iterating for all terms yields

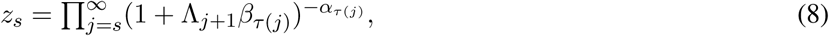

which is an exact expression for *z*_*s*_. As a string of zero incidence values accumulates with time Λ_*j*+1_ → 0 and hence *q*_*j*_ → 1. Consequently, only a finite number of terms in Eq. (8) need to be computed and the initial ones are the most important for evaluating *z*_*s*_.

The posterior mean estimate of *R*_*τ* (*s*)_ is 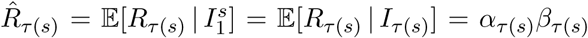 with *I*_*τ* (*s*)_ as the incidence values in the *τ* (*s*) window (the remaining 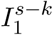 are assumed uninformative [12]). This follows from the Gam distribution and implies a posterior mean incidence prediction 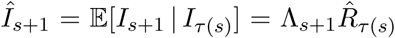 from the NB posterior predictive distribution [22]. Substituting these into Eq. (8) gives:

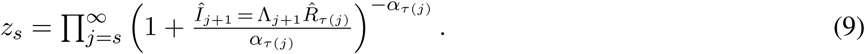

This completes the derivation. Theorem 1, when combined with Eq. (6), provides a new, analytic and event-triggered approach to adjudging when an outbreak has ended. Eq. (9) provides direct and quantifiable insight into what controls the elimination of an epidemic and can be easily computed and updated in real time.

### Understanding the probability of elimination

We dissect and verify the implications of Theorem 1, which provides an exact formula for estimating the probability, *z*_*s*_, that any infectious disease epidemic has been eliminated by time *s*. Eq. (8) formalises the expectation that any decrease in case incidence increases *z*_*s*_. This results because 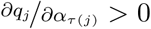 0 for all *α*_*τ*(*j*)_, meaning that *q*_*j*_ is monotonically increasing in *α*_*τ* (*j*)_ and hence *i*_*τ* (*j*)_. As *z*_*s*_ is a product of *q*_*j*_ and every *q*_*j*_ is positive then *z*_*s*_ is also monotonically increasing in all incidence window sums. Consequently, any process that reduces historical or cumulative incidence surely increases the probability of elimination.

The main variable controlling *z*_*s*_ is the average predicted incidence *Î*_*j*+1_ (see Eq. (9)). Reducing either Λ_*j*+1_ or 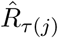 therefore increases our confidence in a declaration made after a fixed time (the time-triggered approach) or, decreases the time of declaration for a fixed confidence (the event-triggered approach). This highlights the two known ways that sustained interventions, e.g. vaccination, social-distancing or quarantine, can help drive an epidemic to extinction. First, such measures explicitly limit *R*_*j*_ and hence 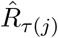, leading to an expected rise in *z*_*s*_ [23]. Second, they may also implicitly reduce the duration of the serial interval, resulting in smaller Λ_*j*+1_ [30].

>Accordingly, under- or over-estimating 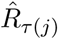 or using incorrectly smaller or larger Λ_*j*+1_ sums induces spurious fluctuations in *z*_*s*_ and promotes premature or delayed declarations, respectively. This insight underlies later analyses, which investigate how surveillance imperfections can modulate the declaration time. Because we cannot reduce either reproduction numbers or serial intervals to arbitrary values of interest (e.g. certain diseases have intrinsically wider serial interval distributions) some epidemics will be innately harder to control and eliminate [31].

Interestingly, while *z*_*s*_ is controlled by mean estimates and predictions, it appears insensitive to the uncertainty around those means, despite its derivation from the posterior distributions of Eq. (1) and Eq. (2). This follows from the inherent data shortage at the tail of an epidemic (there are necessarily many zero incidence points), which likely precludes the inference of higher order statistics [24]. Moreover, when the incidence is small stochastic fluctuations can dominate epidemic dynamics. Consequently, to maximise the reliability of our *z*_*s*_ estimates we recommend using long windows (large *k*) for 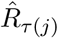. Short windows are more sensitive to recent fluctuations and are more prone to yielding uninformative estimates when many zero incidence points occur [22].

Last, we validate the correctness of our estimated *z*_*s*_ by considering a hypothetical setting in which the true reproduction number, {*R*_*s*_: *s* ≥ 0}, is known without error. This allows us to derive the true (but unknowable) probability of elimination 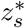 at time *s*, given complete information of the epidemic statistics. Under the renewal model 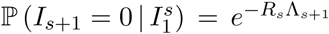. Repeating this process sequentially for future zero infected cases (akin to describing the likelihood of that observation series) gives:

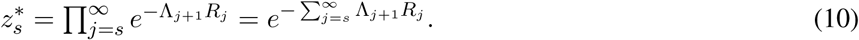

Clearly 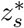 depends on the serial interval distribution and past incidence (through Λ_*j*+1_) and the sequence of reproduction numbers *R*_*j*_, which are the main factors underlying the transmission of the infectious disease.

The true declaration time with confidence *µ*% is then 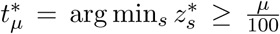 (see Eq. (6)). We can verify our approach to end-of-epidemic declarations if we can prove that *t*_*µ*_ sensibly converges to 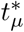. At the limit of *α*_*τ* (*j*)_ → *i*_*τ* (*j*)_ → ∞, the estimated 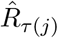 tends to the true *R*_*j*_ because under those conditions the posterior mean estimate coincides with the grouped maximum likelihood estimate of *R*_*j*_, which is unbiased. Applying this limit to *q*_*j*_ in Eq. (9) we find that as 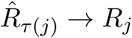:

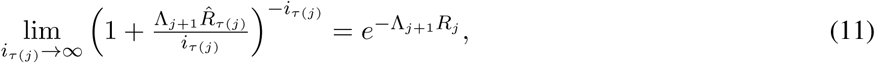

implying that 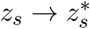, and consequently that 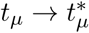.

This asymptotic consistency suggests that *z*_*s*_ and *t*_*µ*_ indeed approximate the true but unknowable probability of elimination 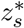 and declaration time 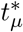. Other end-of-epidemic metrics in the literature have not shown such theoretical justification. We illustrate *z*_*s*_ and 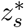 across a simulated and representative incidence curve in Fig. 2. There we find a good correspondence between these probabilities and observe their sensitivity to changes in incidence at the beginning and end of this outbreak. Note that *z*_*s*_ and 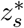 (and hence *t*_*µ*_ and 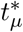) depend on 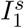 and are more precisely written as 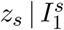 and 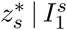. The WHO declaration time, *t*_*WHO*_, which is included for reference, is mostly independent of the shape of 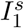 [3], explaining why it provides no confidence guarantee.

### Practical comparisons and verification

We have only validated our approach at an asymptotic limit that is not realistic for elimination i.e. the proof that *z*_*s*_ and *t*_*µ*_ converge to their true counterparts requires infinite incidence. While this proof suggests our formulation is mathematically correct, it does not indicate its performance on actual elimination problems. We now verify out method more practically. We first use simulated data to show that Δ*t*_*µ*_ = *t*_*µ*_ − *t*_0_ and 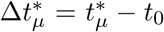 correspond well over several end-of-epidemic problems, where we are far from this limit, and with *t*_0_ as the time of the last observed case. We characterise this via histograms of the error 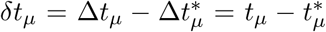, which are given in panels (a)-(c) of Fig. 3. There we present 95% (*µ* = 0.95) declaration time errors over 1000 simulated epidemics with serial interval distributions from the COVID-19 pandemic [32], MERS-CoV in Saudi Arabia [25], Marburg virus in Angola [29] and Measles in Germany [12].

**Fig. 3:**
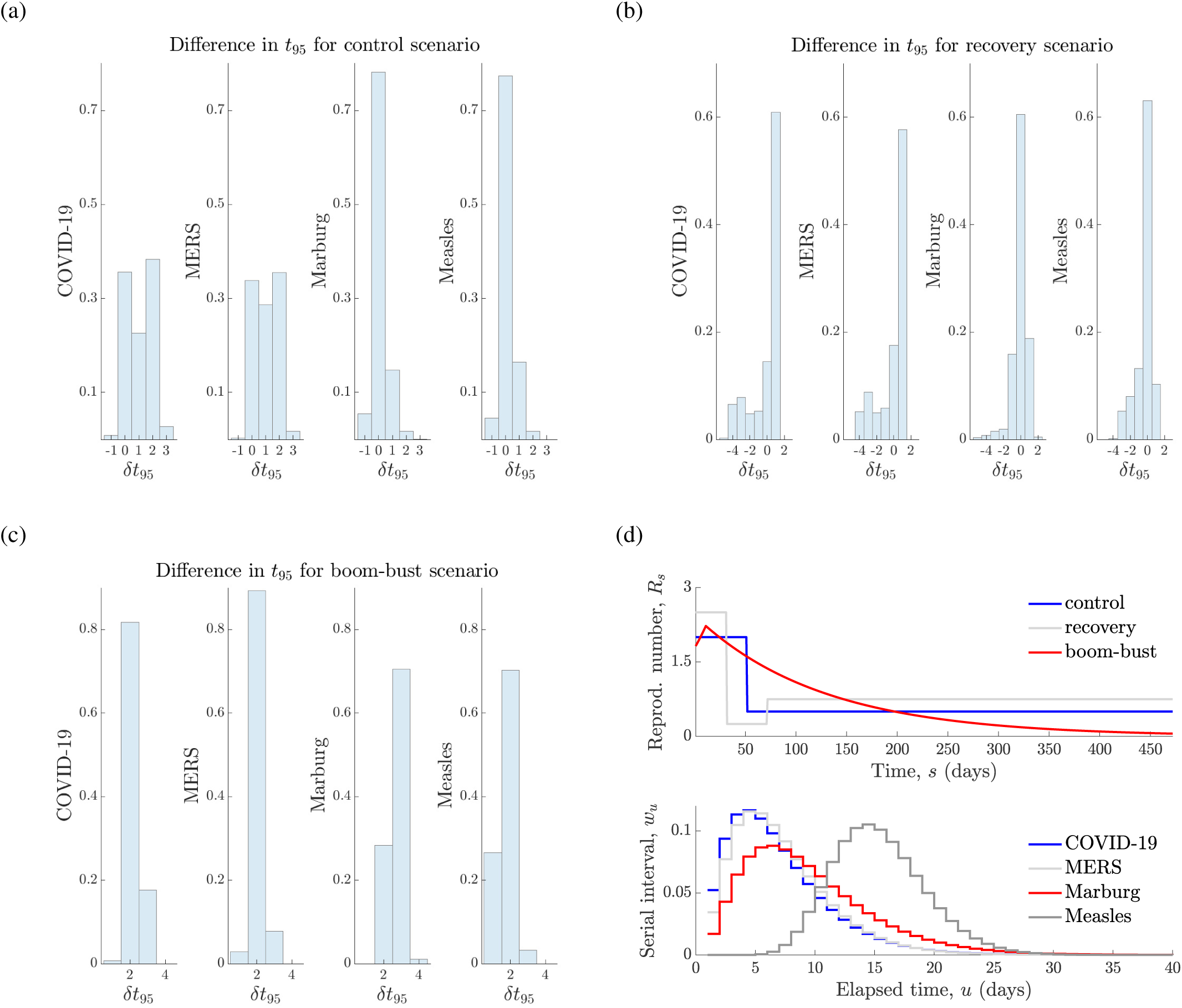
True and estimated declaration times. We simulate 1000 independent incidence curves under various renewal models and provide normalised histograms of the difference between the estimated and true declaration times i.e. 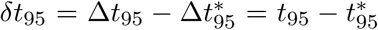. Panels (a)-(c) present these histograms for various infectious diseases under *R*_*s*_ profiles indicating (a) rapidly controlled, (b) recovering and (c) rising and then decaying transmission (boom-bust). The top row of (d) plots the true *R*_*s*_ curves in absolute time, while the bottom row of (d) provides the serial interval distributions of the infectious diseases examined. Generally we find that 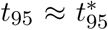 to a reasonable level. The quality of this approximation depends on the variability of the serial interval distribution (see S1 Fig) and the degree of fluctuation in transmission when incidence is small.

We investigate true *R*_*s*_ profiles that describe (a) rapidly controlled, (b) partially recovering and (c) exponentially rising and falling transmission (boom-bust). For each profile we use the renewal model to simulate conditionally independent 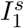 curves and compute 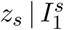 and 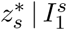 using Eq. (9) and Eq. (10). The declaration time errors then follow as above and from Eq. (6). Panel (d) plots these *R*_*s*_ profiles (top) and the serial interval distributions for each disease (bottom). Generally, we find that *t*_*µ*_ is a good approximation to 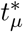, with some error naturally emerging from the difficulty of estimating *R*_*s*_ in conditions where data are necessarily scarce [33]. Our prior distribution over *R*_*τ* (*j*)_ is Gam(1, 5), which is both uninformative and has a large mean of 5.

This error, *δt*_95_, is more prominent for diseases featuring wide serial interval distributions, which are fundamentally more difficult to estimate, due to their dependence on much earlier epidemic dynamics. These simulations also demonstrate why time-triggered approaches can be misleading; they do not adapt to the shape of the specific instance of 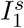 observed. An example of this is given in S1 Fig, where we find that the WHO declaration time Δ*t*_*WHO*_ = *t*_*WHO*_ − *t*_0_ is delayed relative to both the true 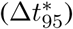 and estimated Δ*t*_95_ event-triggered declaration times, for Ebola virus disease, which has a wide serial interval. Depending on the disease of interest Δ*t*_*WHO*_ could also be premature. The large variability among the possible 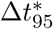 provides a clear visualisation of the non-deterministic nature of epidemic end-points and the need for adaptive metrics with stated confidence.

At present, we have only verified our method under ideal reporting conditions. Practical surveillance is investigated in later sections. We now compare our method to the event-triggered one of [2], which assumes ideal surveillance and models epidemic transmission with a NB branching process that is strictly only valid at the beginning of the outbreak. This notably differs from our renewal model approach and the elimination probabilities derived in [2] are a mathematically conservative approximation to our *z*_*s*_. We compare both methods on MERS-CoV data from South Korea, examined in [2], by running them on a set of bootstrapped incidence curves generated from fitting the model of [2] to that data and compute 95% confidence intervals on the probability of elimination.

Fig. 4 presents our main results with time relative to the last observed case in each bootstrap (Δ*s*) and blue and red curves as the outputs of [2] and our method. While the median 95% relative declaration times (black circles) are close, the approach of [2] yields a delayed declaration. This effect is reduced if we use the lower bound of the *z*_*s*_ curves instead of their median. When *z*_*s*_ is small (which is not practical for defining end-of-epidemic declarations) we find that the methods are less consistent. The WHO declaration time (dark blue) for this epidemic is over one week later than the time proposed by both methods [2]. While our method shows wider uncertainty, the similarity of these intervals suggests that our formulation is robust to moderate model mismatch.

**Fig. 4:**
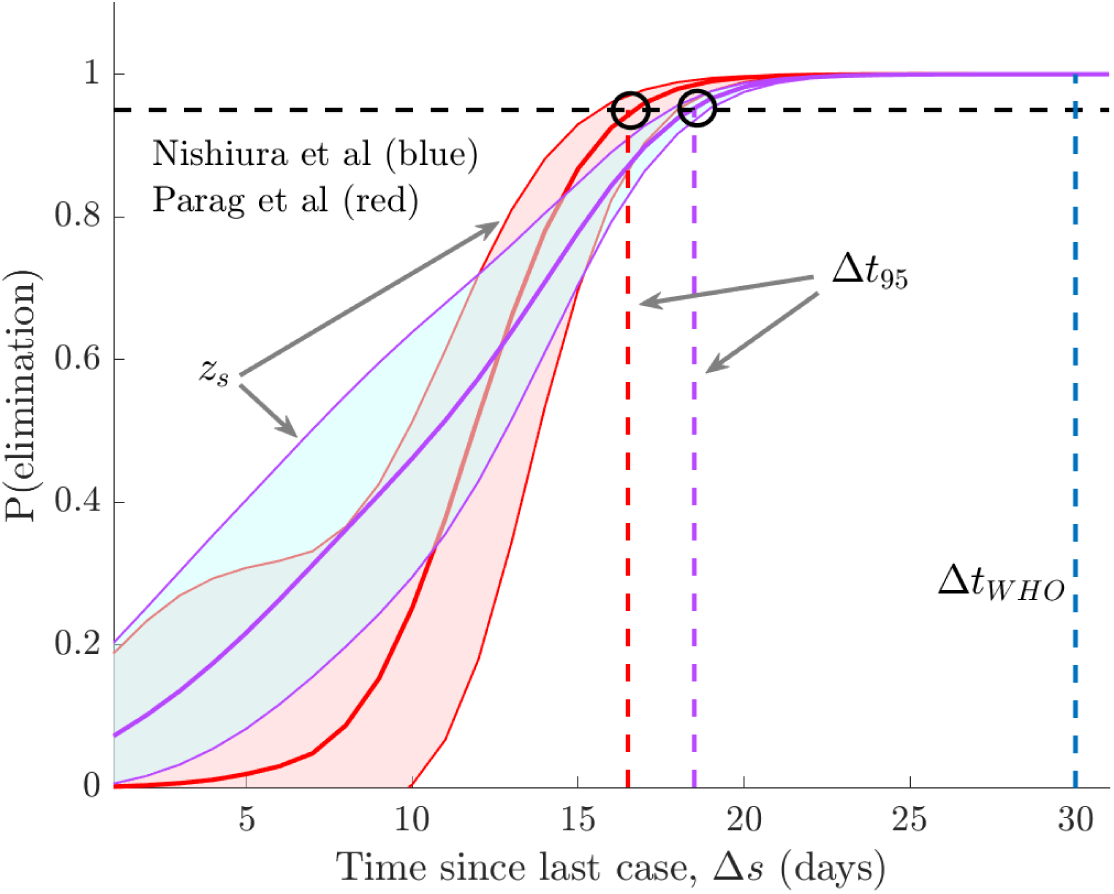
Empirical method comparison. We compare 95% confidence intervals on the elimination probability from [2] (blue) and *z*_*s*_ from Eq. (9) (red) on bootstrapped epidemics based on the MERS-CoV data from South Korea used in [2]. Black circles define the median relative declaration time (Δ*t*_95_) when each method deems the epidemic to be over with 95% confidence (the event trigger). Time is relative to the last observed case in each epidemic bootstrap and the WHO (time-triggered) declaration time (Δ*t*_*WHO*_) is in dark blue.

### Under-reporting leads to premature declarations

Having verified *z*_*s*_ and hence *t*_*µ*_ as reliable and sensible means of assessing the conclusion of an epidemic, we investigate the effect of model mismatch due to imperfect surveillance. We start with case under-reporting, which affects all infectious disease outbreaks to some degree. While previous works have drawn attention to how constant under-reporting can bias end-of-epidemic declarations [6] [7], no analytic results are available. Moreover, the impact of time-varying under-reporting, which models a wide range of more realistic surveillance scenarios [8, 34], remains unstudied. We provide mathematical background for our under-reporting models in Methods.

Fig. 1 illustrates how under-reporting results in only a portion, *N*_*s*_, of the total local cases, *I*_*s*_ being sampled or observed. We use *U*_*s*_ = *I*_*s*_ − *N*_*s*_ ≥ 0 to denote the unreported cases. We investigate two hypotheses or models about the incidence curve, a null one, ℋ_0_, where we assume that the observed cases 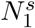 represent all the infected individuals and an alternative hypothesis ℋ_1_, in which the unreported cases 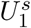 (and hence 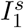) are known and distinguished. The estimated elimination probabilities under both surveillance models are:

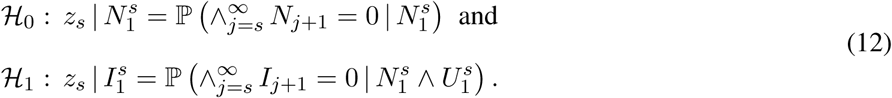

Here ℋ_0_ portrays a naive interpretation of the observed (*N*_*s*_) incidence, while ℋ_1_ indicates ideal surveillance. Intensive and targeted population testing should interpolate between ℋ_0_ and ℋ_1_. We compute 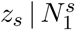 by constructing the sampled total infectiousness 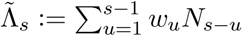 and then applying Theorem 1. This follows because *N*_*s*_ can also be described by a Poisson renewal model (see Methods for details). We therefore find that 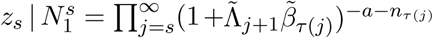 with *n*_*τ* (*j*)_ and 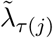 as the sums of *N*_*u*_ and 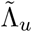 within the *τ* (*j*) window and 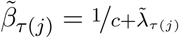 We get 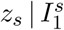 directly from Eq. (8) since this is the perfect surveillance case.

Since *N*_*s*_ ≤ *I*_*s*_ for all *s* then 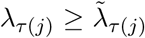 for all *j*, assuming that the same serial interval distribution applies. As a result, 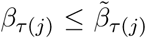, which means that 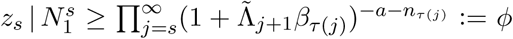. From Eq. (8) we can rewrite 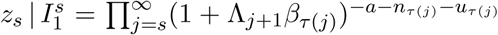 with *u*_*τ* (*j*)_ = *i*_*τ* (*j*)_ − *n*_*τ* (*j*)_ as the total number of unreported cases in the window *τ* (*j*). We examine the ratio of 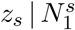 to 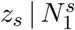, which is at least as large as 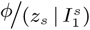. If this ratio is above 1 then the elimination probability is being inflated by imperfect surveillance. We find that 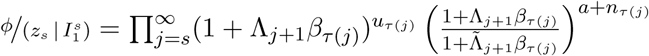. Since 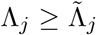 at every *j* (historical numbers of cases are fewer) and the remaining term is always ≥ 1 we do find this inflation and consequently

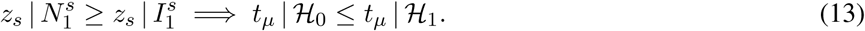

At no point have we assumed any form for the under-reporting fraction, denoted *ρ*_*s*_ at time *s* (see Methods). Our derivation only depends on under-reporting causing smaller (absolute) historical incidence.

Thus any under-reporting, whether constant (i.e. all *ρ*_*s*_ are the same) or time-varying will engender premature or false-positive end-of-epidemic declarations provided *N*_*s*_ is randomly sampled from *I*_*s*_ (so Theorem 1 holds; see Eq. (3)). We highlight this principle by examining a random sampling scheme using empirical SARS 2003 data from Hong Kong [12]. We binomially sample the SARS incidence with random probability *ρ*_*s*_ ∼ Beta(*a, b*). We set *b* = 40 and compute *a* so that the mean sampling fraction 𝔼[*ρ*_*s*_] = *f*_*ρ*_ takes some desired (fixed) value. We investigate various *f*_*ρ*_ and show that premature declarations are guaranteed in (a) and (b) of Fig. 5. The impact of *ρ*_*s*_ is especially large when under-reporting leads to early but false sequences of 0 cases, which is additional to the bias from Eq. (13). We present results in absolute time to showcase this effect.

**Fig. 5:**
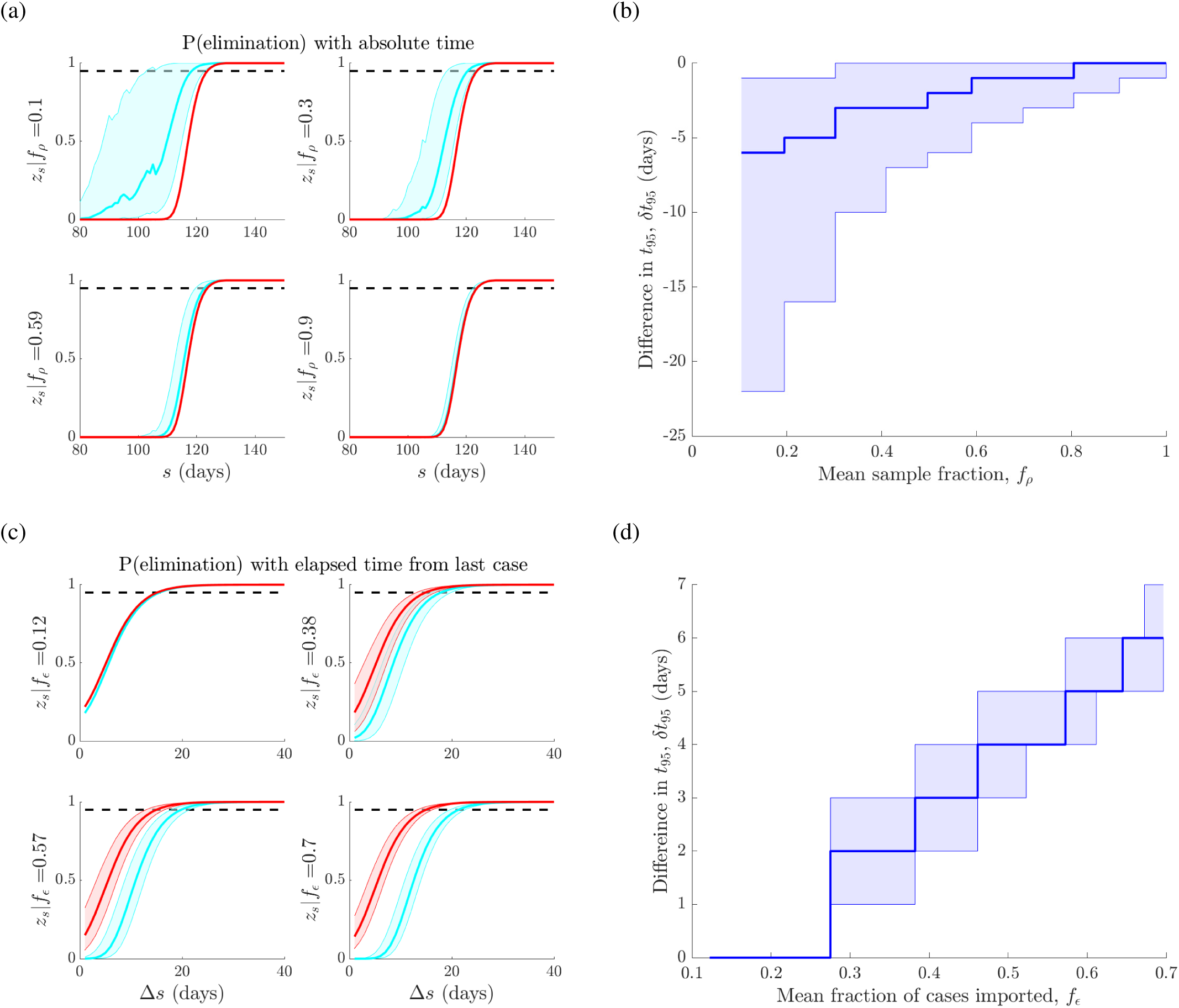
Case under-reporting and importation lead to premature and delayed declarations respectively. In (a) and (b) we binomially sample an empirical SARS 2003 incidence curve from Hong Kong with reporting probabilities drawn from a beta distribution with mean *f*_*ρ*_. In (a) we plot the elimination probability *z*_*s*_ when surveillance is ideal i.e. there is no underreporting (red) versus when the under-reporting is unknown (blue). The difference in the 95% declaration times, denoted *δt*_95_, from these curves is in (b). As *f*_*ρ*_ increases we are more likely to declare too early. In (c) and (d) we consider an empirical MERS-CoV 2014-5 incidence curve from Saudi Arabia with local and imported cases. We increase the mean fraction of imported cases to *f*_*ϵ*_ by adding Poisson imports with mean *ϵ* and in (c) compute *z*_*s*_ with (red) and without (blue) accounting for the difference between imports and local cases. The change in *t*_95_ is given in (d). As *E* and hence *f*_*ϵ*_ increase later declarations become more likely. We repeat our sampling or importation procedure 1000 times to obtain confidence intervals in (a)–(d). As *f*_*ϵ*_ → 0 or *f*_*ρ*_ → 1 we attain the ideal surveillance scenarios of no unreported or imported cases.

### Importation results in late declarations

The influence of imported cases on end-of-epidemic declarations, to our knowledge, has not been investigated in the literature. Repeated importations or migrations of infected cases are a common means of seeding and re-seeding local infectious epidemics. Failing to ascertain which cases are local or imported can significantly change our perception of transmission [9]. We assume that *I*_*s*_ is the total count of local cases in our region of interest but that at time *s* there are also *M*_*s*_ imported cases that have migrated from neighbouring regions. The total number of infected cases observed is *C*_*s*_ = *I*_*s*_ + *M*_*s*_ as displayed in Fig. 1. We provide mathematical background on how importations are included within the renewal framework in Methods. We consider two hypotheses about our observed incidence data that reflect real epidemic scenarios.

Under the null hypothesis, ℋ_0_, we assume that all cases are local and so we cannot disaggregate the components of *C*_*s*_. The alternative, ℋ_1_, assumes perfect surveillance. Imported cases are distinguished from local ones under ℋ_1_ and their differing impact considered. The relevant elimination probabilities for each model are

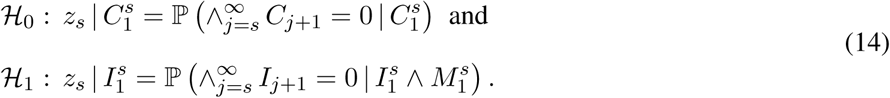

Since ℋ_0_ deems all cases local, it models *C*_*s*_ as a renewal process with total infectiousness 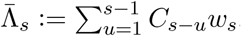. Thus we use Theorem 1 to obtain the *j*^th^ factor of 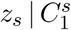 as 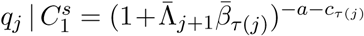 with 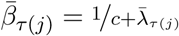. Here *c*_*τ* (*j*)_ and 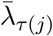 are sums of *C*_*u*_ and 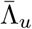 over window *τ* (*j*).

Under ℋ_1_ the imported cases are distinguished but all cases still contribute to ongoing local transmission [9, 28]. Consequently, *I*_*s*_ still adheres to a renewal transmission process and Theorem 1 yields the *j*^th^ factor of 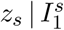 as 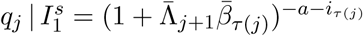. We compare 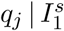 with 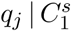 directly to easily prove that

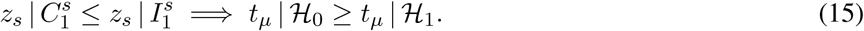

Not accounting for migrations shrinks the elimination probability leading to false-negative or unnecessarily late declarations. This result makes no assumption on the dynamics for importation other than it possesses Poisson noise (so Theorem 1 is valid for *C*_*s*_) and so holds quite generally (see Methods for further details).

We illustrate this phenomenon using empirical MERS-CoV data from Saudi Arabia [25] in (c) and (d) of Fig. 5. Here repeated importations occur as zoonotic camel to human transmissions. We show the increasing effect of importation by adding further (artificial) imports via a Poisson noise variable with mean *ϵ* (see Eq. (4)). The mean fraction of imported to total cases across the incidence curve is then *f*_*ϵ*_. In Fig. 5 we see that larger *ϵ* promotes increasingly later declaration times. In Fig. 5 we do not add any imports beyond the time of the last local case. If imports do come after this case, and seed no further local infections, which is likely for epidemics with large heterogeneity, then the *t*_0_ assumed under ℋ_0_ will be later, and further exacerbate the bias from importation.

## Discussion

Understanding and predicting the temporal dynamics of infectious disease transmission in real time is crucial to controlling existing epidemics and to thwarting future resurgences of those outbreaks, once controlled [21]. To achieve this understanding it is necessary to characterise and study the infectious disease throughout its lifetime. While many works have focussed on the growth, peak and controlled phases of epidemics (see Fig. 2), relatively less research has examined how the tail of the outbreak shapes the kinetics of its elimination. For example, while much is known about how the basic and effective reproduction numbers influence the growth rate, peak size and controllability of an epidemic [4, 35], the relationship between these numbers and the waiting time to epidemic elimination or extinction is still largely unexplored.

However, this relationship has important implications for public health policy. Knowing when to relax non-pharmaceutical interventions, such as social distancing or lockdowns, can be essential to effectively managing and mitigating the financial and social disruption caused by an outbreak as well as to safeguarding populations from the risk of future waves of the disease [1, 2]. The ongoing COVID-19 pandemic for instance, which in some countries such as New Zealand entered a prolonged period of near-elimination before resurgence occurred [33], provides a current and important example where this question might soon become urgent.

Existing WHO guidance on deciding when an outbreak can be safely declared over takes a time-triggered approach. This means a fixed waiting time from the last observed case, usually based on the incubation period of the disease, is adopted [3]. While this approach is easy to follow, it does not change informatively between outbreaks of the same disease, even if the patterns of transmission are very different and cannot provide a measure of the reliability of this suggested declaration time. The few existing studies that have investigated this waiting-time problem [2, 6, 7] have all converged to what is known as an event-triggered solution in control theory [13].

Event-triggered decision-making has been shown to be more effective than acting at deterministic or fixed times for a range of problems including several involving the optimising of waiting or stopping times [14, 15, 16, 17]. Moreover, because it directly couples decision making to observables of interest (in our case the incidence curve), it can better adapt or respond to changes in dynamics. Here we have attempted to build upon these realisations to better characterise the relationship between epidemic transmission and elimination. Specifically, we focussed on computing the probability at time *s, z*_*s*_, that the total future incidence of the epidemic is zero.

This probability is directly responsible for determining how quickly an epidemic will end. In fact, if an outbreak is defined as surviving if it can propagate at least 1 future infection then 1−*z*_*s*_ is precisely its survival function and is therefore rigorously linked to the future risk of cases. By taking a renewal process approach, we were able to derive an analytic and real-time measure of *z*_*s*_ that explicitly depends on up-to-date estimates of the effective reproduction number (see Eq. (9)). This result formed the main theorem of this paper and provided a clear and easily-computed link between epidemic transmission and elimination. To our knowledge, no previous work has directly obtained *z*_*s*_. Specifically, [2] computed a simpler and more conservative quantity while [6] and [7] approximated something similar via simulation, and so cannot provide real-time formulae. The event-trigger for declaring an outbreak over with *µ*% confidence is then the first time that *z*_*s*_ crosses a threshold of 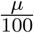.

To validate the correctness of our approach we considered several comparisons. We proved mathematically that our formulae recover the true elimination probability and event trigger given perfect knowledge of the epidemic. This provided theoretical justification for our approach (Eq. (11)). We verified practical performance by benchmarking our method against the known (true) declaration times from simulated outbreaks of several infectious diseases (Fig. 3) and on empirical data by directly comparing to [2] (Fig. 4). We found that our method generated sensible and reasonably accurate estimates, given the fundamental difficulties of inferring *R*_*s*_ at low incidence. Integrating our method with newly developing approaches that improve on *R*_*s*_ estimates in these low data conditions [33], should further enhance performance and forms part of our future work.

Fig. 3, Fig. 4 and S1 Fig also explained why time-triggered methods, such as the existing WHO guidelines, can be unreliable or deceptive. Replicate epidemics driven by the same time-series of reproduction numbers can engender significantly different relative declaration times Δ*t*_95_. This variability exists even if *R*_*s*_ is known perfectly (i.e. when we have 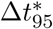). As no single, fixed time can reasonably approximate this distribution, time-triggered approaches are necessarily performance limited. Moreover, we can never guarantee the confidence in such a declaration because *z*_*s*_ and 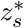 also vary considerably for epidemics of the same disease even under identical transmission dynamics. These issues will only worsen with the additional noise deriving from non-ideal surveillance.

Exploring non-ideal surveillance noise and rigorously assessing its impact on the tail dynamics of epidemics was the main motivation for developing our method. Consequently, we investigated two prevalent and potentially dominant sources of noise in surveillance – unreported and imported cases [9, 26]. While both [6] and [7] looked at the effect of constant under-reporting on declarations, general insight into the more realistic time-varying case is lacking. Further, the influence of importation on the epidemic tail has, to our knowledge, not yet been examined. By adapting *z*_*s*_ to various surveillance hypotheses we proved two key results and developed a flexible framework for incorporating and analysing the influence of other related noise sources.

First, we showed that any type of random under-reporting will precipitate early declarations, which worsen as the fraction of unreported cases increases (Eq. (13)). Second, we found that any random importation process will lead to late declarations that become more delayed as the fraction of imports increase (Eq. (15)). Moreover, under-reporting and importation processes can respectively, cause falsely early and late starts (i.e. *t*_0_ in our notation) to the sequence of zero incidence days that are used to determine declaration times, thus exacerbating the bias from each noise source. We illustrated the biases of both unreported and imported cases using empirical data (Fig. 5), clarifying how the epidemic tail is sensitive to these imperfections in the collection or reporting of incidence data. The theoretical framework we employed to reveal these biases can also help generate insight into other noise sources and surveillance hypotheses. It provides a scheme for investigating case misidentification, asymptomatic transmission and reporting delays, among others. The first occurs when cases of a co-circulating diseases are misattributed to the disease of interest due to overlapping symptoms and is common among influenza-like illnesses [8]. The disease of interest is then effectively over-reported, which may be modelled as a false importation process with *M*_*s*_ as the over-reported cases in Eq. (4), but past *M*_*s*_ counts do not contribute to *I*_*s*_ (and so are not in its total infectiousness term). It then follows that declaration times will be delayed.

Asymptomatic transmission and reporting delays are effectively types of under-reporting. In the first, the cases observed at any time represent only the symptomatic fraction of actual infections. Consequently, a formulation similar to Eq. (3) applies, with variations depending on whether the asymptomatic proportion has the same or a different serial interval distribution [36]. The result is that end-of-epidemic declarations that do not account for asymptomatic transmission will be early. Reporting delays act as time-varying under-reporting fractions, which especially degrade the more recent case days [10]. While the model required is more involved than Eq. (3), since the declaration times largely depend on cumulative case counts, they are also likely to be premature.

While our method presents a clean framework for estimating the lifetime of an epidemic and investigating surveillance noise sources, it has several limitations. It commonly assumes that the serial interval distribution is known [12]. However, if surveillance is poor and changes to the serial interval (e.g. contractions due to interventions [30]) are not measured or included in computing *z*_*s*_ then declaration times might be biased. Moreover, we neglect transmission heterogeneity, are necessarily hindered by the difficulty of estimating reproduction numbers at low incidence and do not consider interactions among noise sources. While these factors could limit the accuracy of our predicted declaration times, many can be accommodated as future extensions. We can incorporate heterogeneity by using negative binomial renewal models [1], improve on low incidence estimates by capitalising on specialised methods [33] and extend the models in Fig. 1 to examine mixed noise types.

A key contribution of this work has been clarifying and highlighting how realistic imperfections in the collection or reporting of incidence data can significantly influence and bias the tail dynamics of an epidemic. Heightened surveillance should therefore be sustained even in periods of negligible incidence. Intensive testing and tracing is especially essential as it provides a means of measuring and compensating for case under-reporting, which we found to be among the strongest sources of bias. Maintaining good quality screening and geodata is also important since having accurate case origins can prevent misidentification, which is a main cause of unknown or unrecognised imports. These sentiments echo many issues currently being faced across the COVID-19 pandemic [19, 37].

Real-time assessments of epidemic dynamics are crucial for understanding and aptly responding to unfolding epidemics [21]. We hope that the analytic approach developed here will serve as a useful tool for gaining ongoing insight into the tail dynamics of an outbreak, motivate the adoption of more event-triggered decision making and provide clear impetus for improving and sustaining surveillance across all phases of an epidemic. Our method is available at https://github.com/kpzoo/End-of-epidemic-declarations. Our future work aims to develop this tool from its current form as a passive means of understanding and uncovering biases to an approach that can actively infuse additional data streams (e.g. case ascertainment ratios) to compensate for these biases in end-of-epidemic declarations.

## Data Availability

All data and code are freely available at https://github.com/kpzoo/End-of-epidemic-declarations

## Acknowledgments

KVP and CAD acknowledge joint centre funding from the UK Medical Research Council and Department for International Development under grant reference MR/R015600/1. RNT thanks Christ Church (Oxford) for funding via a Junior Research Fellowship. CAD thanks the UK National Institute for Health Research Health Protection Research Unit (NIHR HPRU) in Modelling Methodology at Imperial College London in partnership with Public Health England (PHE) for funding (grant HPRU-2012–10080). The funders had no role in study design, data collection and analysis, decision to publish, or preparation of the manuscript.

## Author Contributions

Conceptualization: KVP and RNT. Formal analysis, investigation, methodology, project administration, software, visualisation and writing (original draft preparation): KVP. Validation: KVP, RJ, RNT and CAD. Writing (review and editing): KVP, RNT, RJ and CAD.

## Supporting Information

**Fig. S1:**
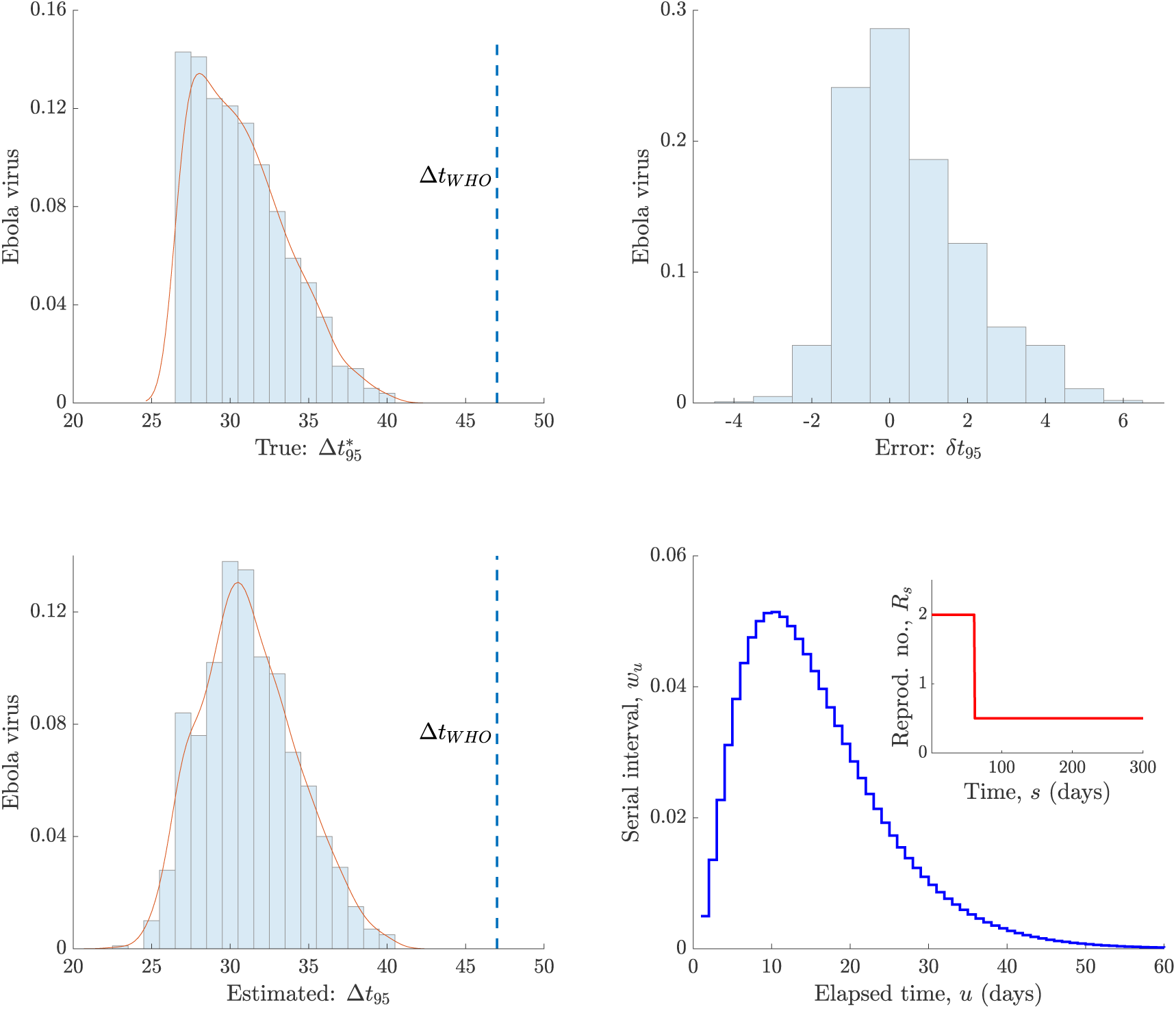
Event and time-triggered declarations. We compare 95% event-triggered declaration times to the WHO time-triggered equivalent for Ebola virus disease over 1000 simulated epidemics. Left panels show the true 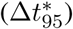 and estimated (Δ*t*_95_) declaration times (based on Eq. (10) and Eq. (9)) relative to the time of the last observed case. The significant variability in both, which reflects the different shapes of possible epidemic curves with the same reproduction number profile (*R*_*s*_) indicates why time-triggered approaches such as the WHO one [3] (Δ*t*_*WHO*_, which is based on 42 days plus the time to recovery) can be insufficient. The error between the true and estimated times (*δt*_95_) and the serial interval and reproduction number profile used are shown in the right panels.

## Footnotes

1 Strictly, it is the time elapsed since the last case has recovered or died. However, as this additionally delay is not informative, it does not invalidate or alter any of the results or statements in this paper and so we speak in terms of the last case time for simplicity.

## References

1. Lee H, Nishiura H. Sexual transmission and the probability of an end of the Ebola virus disease epidemic. J Theor Biol. 2019;471:1–12.

2. Nishiura H, Miyamatsu Y, Mizumoto K. Objective determination of end of MERS outbreak, South Korea. Emerg Infect Dis. 2016;22:146–8.

3. WHO. WHO recommended criteria for declaring the end of the Ebola virus disease outbreak; 2020. Available from: https://www.who.int/who-documents-detail/who-recommended-criteria-for-declaring-the-end-of-the-ebola-virus-disease-outbreak.

4. Wallinga J, Lipsitch M. How generation intervals shape the relationship between growth rates and reproductive numbers. Proc R Soc B. 2007;274:599–604.

5. Fraser C. Estimating Individual and Household Reproduction Numbers in an Emerging Epidemic. PLOS One. 2007;8:e758.

6. Thompson R, Morgan O, Jalave K. Rigorous surveillance is necessary for high confidence in end-of-outbreak declarations for Ebola and other infectious diseases. Phil Trans R Soc B. 2019;374:20180431.

7. Djaafara B, Imai N, Hamblion E, et al. A quantitative framework to define the end of an outbreak: application to Ebola Virus Disease. medRxiv. 2020;(20024042).

8. White L, Pagano M. Reporting errors in infectious disease outbreaks, with an application to Pandemic Influenza A/H1N1. Epidemiol Perspec Innov. 2010;7(12).

9. Churcher T, Cohen J, Ntshalintshali N, et al. Measuring the path toward malaria elimination. Science. 2014;344(6189):1230–32.

10. Yan P, Chowell G. Quantitative Methods for Investigating Infectious Disease Outbreaks. vol. 70 of Texts in Applied Mathematics. Cham, Switzerland: Springer; 2019.

11. Fraser C, Cummings D, Klinkenberg D, et al. Influenza Transmission in Households During the 1918 Pandemic. Am J Epidemiol. 2011;174(5):505–14.

12. Cori A, Ferguson N, Fraser C, et al. A New Framework and Software to Estimate Time-Varying Reproduction Numbers During Epidemics. Am J Epidemiol. 2013;178(9):1505–12.

13. Astrom K, Bernhardsson B. Comparison of periodic and event based sampling for first order systems. Proc IFAC World Conf. 1999:301–6.

14. Parag K. On signalling and estimation limits for molecular birth-processes. J Theor Biol. 2019;480:262–73.

15. Rabi M, Moustakides G, Baras J. Adaptive Sampling for Linear State Estimation. SIAM Journal of Control and Optimization. 2012;50(2):672–702.

16. Parag K, Vinnicombe G. Point Process Analysis of Noise in Early Invertebrate Vision. PLOS Comput Biol. 2017;13(10):e1005687.

17. Lemmon M. Event-Triggered Feedback in Control, Estimation, and Optimization. vol. 406 of Networked Control Systems. London: Springer; 2010. p. 293–358.

18. Bhatia S, Cori A, Parag K, et al.. Short-term forecasts of COVID-19 deaths in multiple countries.; 2020. Available from: https://mrc-ide.github.io/covid19-short-term-forecasts.

19. Pybus O, Rambaut A, du Plessis L, Zarebski A, et al.. Preliminary analysis of SARS-CoV-2 importation & establishment of UK transmission lineages; 2020. Available from: https://virological.org/t/preliminary-analysis-of-sars-cov-2-importation-establishment-of-uk-transmission-lineages [cited 13 June 2020].

20. Thompson R, Hollingsworth D, Isham V, et al. Key questions for modelling COVID-19 exit strategies. Proc R Soc B. 2020;287(1932):20201405.

21. Cauchemez S, Boelle P, Thomas G, et al. Estimating in Real Time the Efficacy of Measures to Control Emerging Communicable Diseases. Am J Epidemiol. 2006;164(6):591–7.

22. Parag K, Donnelly C. Using information theory to optimise epidemic models for real-time prediction and estimation. PLOS Comput Biol. 2020;16(7):e1007990.

23. De Serres G, Gay N, Farrington P. Epidemiology of Transmissible Diseases after Elimination. Am J Epidemiol. 2000;151(11).

24. Parag K, Donnelly C. Adaptive Estimation for Epidemic Renewal and Phylogenetic Skyline Models. Syst Biol. 2020;(syaa035).

25. Thompson R, Stockwin J, van Gaalen R, et al. Improved inference of time-varying reproduction numbers during infectious disease outbreaks. Epidemics. 2019;29:100356.

26. Azmon A, Faes C, Hens N. On the estimation of the reproduction number based on misreported epidemic data. Stats Med. 2014;33:1176–92.

27. Raikov D. On the decomposition of Poisson laws. Dokl Acad Sci URSS. 1937;14:9–11.

28. Roberts M, Nishiura H. Early Estimation of the Reproduction Number in the Presence of Imported Cases: Pandemic Influenza H1N1- 2009 in New Zealand. PLOS One. 2011;6(5):e17835.

29. Van Kerkhove M, Bento A, Mills H, et al. A review of epidemiological parameters from Ebola outbreaks to inform early public health decision-making. Sci Data. 2015;2:150019.

30. Ali S, Wang L, Lau E, et al. Serial interval of SARS-CoV-2 was shortened over time by nonpharmaceutical interventions. Science. 2020;369(6507):1106–9.

31. White L, Pagano M. A likelihood-based method for real-time estimation of the serial interval and reproductive number of an epidemic. Stats Med. 2008;27:2999–3016.

32. Ferguson N, Laydon D, Nedjati-Gilani G, et al. Impact of non-pharmaceutical interventions (NPIs) to reduce COVID- 19 mortality and healthcare demand. Imperial College London; 2020.

33. Parag K. Improved estimation of time-varying reproduction numbers at low case incidence and between epidemic waves. medRxiv. 2020;2020.09.14.20194589.

34. Parag K, du Plessis L, Pybus O. Jointly inferring the dynamics of population size and sampling intensity from molecular sequences. Mol Biol Evol. 2020;37(8):2414–29.

35. Brauer F, van den Driessche P, Wu J, editors. Mathematical Epidemiology. Lecture Notes in Mathematics. Berlin, Germany: Springer-Verlag; 2008.

36. Park S, Cornforth D, Dushoff J, et al. The time scale of asymptomatic transmission affects estimates of epidemic potential in the COVID-19 outbreak. Epidemics. 2020;31:100392.

37. Li R, Pei S, Chen B, et al. Substantial undocumented infection facilitates the rapid dissemination of novel coronavirus (SARS-CoV-2). Science. 2020;368(6490):489–93.

